# Derivation, validation, and clinical relevance of a pediatric sepsis phenotype with persistent hypoxemia and shock

**DOI:** 10.1101/2022.12.23.22283912

**Authors:** L. Nelson Sanchez-Pinto, Tellen D. Bennett, Emily Stroup, Yuan Luo, Mihir Atreya, Juliane Bubeck Wardenburg, Grace Chong, Alon Geva, E. Vincent S. Faustino, Reid W. Farris, Mark Hall, Colin Rogerson, Sareen Shah, Scott L. Weiss, Robinder G. Khemani

## Abstract

**Background:** Untangling the heterogeneity of sepsis in children and identifying clinically relevant phenotypes could lead to the development of enrichment strategies and targeted therapies. In this study, our aim was to analyze the organ dysfunction-based trajectories of children with sepsis-associated multiple organ dysfunction syndrome (MODS) to identify and characterize reproducible and clinically relevant sepsis phenotypes using a data-driven approach.

**Methods:** We collected data from patients admitted with suspected infections to 13 pediatric intensive care units (PICUs) in the U.S. between 2012-2018. We used subgraph-augmented nonnegative matrix factorization to identify candidate trajectory-driven phenotypes based on the type, severity, and progression of organ dysfunction in the first 72 hours of PICU admission. We analyzed the candidate phenotypes to determine reproducibility as well as prognostic, therapeutic, and biological relevance.

**Results:** Overall, 38,732 children had suspected infection, of which 15,246 (39.4%) had sepsis-associated MODS. Amongst patients with sepsis-associated MODS, 1,537 (10.1%) died in the hospital. We identified an organ dysfunction trajectory-based phenotype (which we termed *persistent hypoxemia and shock*) that was highly reproducible, had features of systemic inflammation and coagulopathy, and was independently associated with higher mortality. In a propensity score matched analysis, patients with the *persistent hypoxemia and shock* phenotype appeared to have a higher likelihood to benefit from adjuvant therapy with hydrocortisone and albumin than other patients. When compared to other high-risk clinical syndromes, the *persistent hypoxemia and shock* phenotype only overlapped with 50 to 60% of patients with septic shock, those with moderate-to-severe pediatric acute respiratory distress syndrome, or those in the top tertile of organ dysfunction burden, suggesting that it represents a distinct clinical phenotype of sepsis-associated MODS with a disproportionately high risk of mortality.

**Conclusions:** We derived and validated the *persistent hypoxemia and shock* phenotype, a trajectory-based organ dysfunction phenotype which is highly reproducible, clinically relevant, and associated with heterogeneity of treatment effect to common adjuvant therapies. Further validation is warranted. Future studies are needed to validate this phenotype, assess whether it can be predicted earlier in the course, study possible biological mechanisms underlying it, and investigate candidate therapeutic targets.

## INTRODUCTION

Sepsis is the most common cause of multiple organ dysfunction syndrome (MODS) in children, which in turn frequently leads to death [1–4]. Although respiratory failure is the leading organ dysfunction associated with pediatric sepsis and sepsis-related deaths [5,6], significant heterogeneity exists in the clinical presentation and underlying pathobiology of children with sepsis [7,8]. Untangling the clinical and biological heterogeneity in sepsis and acute respiratory distress syndrome (ARDS) has been identified by the American Thoracic Society as a major research priority in the path towards precision medicine in critical care [9].

A major challenge for untangling the heterogeneity of sepsis stems from the reality that the pathobiology and clinical state of septic patients is dynamic [10,11]. Oftentimes, single time-points (such as ICU admission or onset of disease) are used to investigate this heterogeneity, but this approach fails to capture the critical importance of the evolving and dynamic nature of sepsis [12,13]. Data from electronic health records (EHRs) provides an attractive opportunity to perform data-driven analyses that incorporate the dimension of time by leveraging the granular, longitudinal clinical data from thousands of patients [10,14]. Indeed, EHR data from adults with sepsis and children with MODS has been used to uncover subgroups of patients with similar clinical characteristics and trajectories (often called ‘phenotypes’ or ‘subphenotypes’) that have been associated with clinically relevant findings, such as distinct cytokine profiles or heterogeneity of treatment effect to interventions like balanced fluids or hydrocortisone administration [3,15,16]. However, no prior study has used this approach to uncover trajectory-based phenotypes in a large cohort of children with sepsis-associated MODS.

In this study, we aimed to derive and validate clinically relevant sepsis phenotypes using a data-driven approach to analyze organ dysfunction trajectories in a large, multicenter cohort of children with sepsis-associated MODS. We hypothesized that we would identify one or more reproducible sepsis phenotypes based on the trajectories of organ dysfunctions in the acute phase of critical illness and that patients with similar trajectories would have distinct risk factors, biochemical characteristics, response to adjuvant therapies, and clinical outcomes.

## METHODS

### Study design and patient population

This was a retrospective, multicenter, observational cohort study of children 0 to 18 years old admitted to one of 13 participating U.S. pediatric intensive care units (PICUs) in the six-year period between January 1, 2012 and January 1, 2018. Data for patients who had a confirmed or suspected infection (i.e., received systemic antimicrobials and microbiological testing in the +/-24 hour time-window after the first admission to the PICU) were extracted from the EHRs of the participating institutions using structured queries and underwent quality checks for conformity, completeness, and plausibility using Kahn’s framework [17]. Data with potential quality issues were flagged and specific queries submitted back to the sites for clarification or re-extraction. Validation of clinical values was assessed based on clinical standards and verification was performed by comparing the distribution of values and count of variables across sites [17]. The institutional review board (IRB) at Ann & Robert H. Lurie Children’s Hospital of Chicago served as the central IRB for this study. The reporting of this observational cohort study was performed using the Strengthening the Reporting of Observational Studies in Epidemiology (STROBE) reporting guideline [18].

We measured the type, severity, and change in organ dysfunction using the six subscores of the pediatric Sequential Organ Failure Assessment (pSOFA) score [1]. The pSOFA scores offers several advantages for this type of analysis, including a normalized scale that has been previously validated and shown to discriminate poor outcomes in critically ill children early in the course better than other scores, as well as coverage of six key organ dysfunctions in sepsis: neurologic, cardiovascular, respiratory, hepatic, renal, and coagulation [1,19]. The pSOFA subscores were calculated for each 24-hour period between PICU admission and 72 hours, which was used as a proxy for the acute phase of illness [14,20]. Individual pSOFA subscores were carried forward until they were remeasured or the patient died; otherwise, if completely missing they were assumed to be normal and the corresponding subscore was assigned a 0, as previously performed [21,22]. We defined patients with MODS as those with a pSOFA subscore of ≥2 in ≥2 organ systems [3]. For the unsupervised analysis, we only included patients with sepsis-associated MODS, which were defined as those with confirmed or suspected infections on admission and MODS within 72 hours. Patients with immunocompromised status were defined as those with a malignancy and/or transplant comorbidity. The comorbidities of malignancy, transplantation, and technology dependence were based on the classification system developed by Feudtner et al. [3,23,24]. Severity of illness on admission was measured using the Pediatric Risk of Mortality (PRISM) III score [25]. The vasoactive-inotropic score was calculated to determine the degree of shock [26]. The primary outcome was in-hospital mortality and the secondary outcome was persistent MODS on day 7 after PICU admission (including patients who died in the first week) [3].

### Trajectory-based phenotyping

Patients were split into three sets based on site, resulting in one derivation set (data from 7 sites) and two external validation sets (3 sites each), which is consistent with geographic-based external validation, an approach recommended by established guidelines for clinical model development and validation [27]. We first used subgraph-augmented nonnegative matrix factorization (SANMF), an unsupervised trajectory modeling approach, to derive a set of organ dysfunction trajectory-based groups as candidate phenotypes in the derivation set, as previously performed by our group in children with MODS [3]. The SANMF analysis consisted of two parts: (a) subgraph mining, in which the individual trajectories of each of the six pSOFA subscores over the first 72 hours were analyzed to determine common patterns of severity and change of the individual organ dysfunctions across patients; and (b) nonnegative matrix factorization, in which patients were grouped into candidate phenotypes based on similarity in the frequency of the subgraphs. The validation and clinical relevance of these candidate phenotypes was then assessed across three dimensions based on an established framework described by DeMerle et al. [28]. Briefly, the validation of the candidate phenotypes was performed across three dimensions: (i) reproducibility of the candidate phenotypes in the first validation set using the SANMF mixture coefficient matrix to assess external validity; (ii) reproducibility of the candidate phenotypes using different unsupervised trajectory modeling methods in the derivation and first validation set (using repeated measures latent class analysis [RM-LCA] and group-based trajectory modeling [GBTM]) to assess statistical approach validity; and (iii) reproducibility of the candidate phenotypes in the second external validation set using a Random Forest classifier to assess whether the SANMF-based phenotypes groups can be accurately predicted in new datasets. The clinical relevance of the candidate phenotypes was also assessed across three dimensions: (i) prognostic relevance based on the independent association with outcomes; (ii) therapeutic relevance based on the association with response to two adjuvant therapies (intravenous hydrocortisone and albumin); and (iii) pathobiological relevance based on association with biochemical and clinical features linked to plausible disease mechanisms and other clinical syndromes. The adjuvant therapies studied (hydrocortisone and albumin) were chosen *a priori* because they are commonly used in sepsis and have plausible mechanisms of action, but have conflicting evidence regarding their efficacy [5].

Additional details on the Methods are included in the Online Data Supplement.

### Statistical Analysis

Categorical variables were compared using the chi-squared test and continuous variables using the Kruskal-Wallis test. Agreement between models was assessed using Fleiss’ Kappa. Mixed effects logistic regression was used to adjust for confounders when comparing the association of candidate phenotypes with outcomes. The random effect was based on the study site and the fixed effects included age, immunocompromised status, and severity of organ dysfunction in the first 72 hours using the mean pSOFA score. Propensity score matching was used to account for selection bias in the response to treatment analysis. The propensity score to receive either of the studied treatments in the first 24 hours of admission was based on the risk factors that were significantly associated with a higher likelihood of receiving the treatment. To limit the influence of survival bias on whether a patient received or did not receive a treatment, only patients alive for at least 24 hours were included in the propensity matched analysis. A sensitivity analysis of the therapeutic relevance of phenotype membership in all patients was performed using inverse probability treatment weighting (IPTW). Survival analysis to 28 days was performed using Kaplan-Meier curves and Cox regression analysis. Statistical significance was set at p<0.05. Data were analyzed using R version 4.0 (R Project for Statistical Computing, Vienna, Austria) and STATA, version 16 (Stata Corp LLC, College Station, TX).

## RESULTS

### Study population

There were 38,732 children with suspected or confirmed infection on admission to the PICU. Of those, 15,246 (39.4%) had sepsis-associated MODS, of whom 1,537 (10.1%) died in the hospital. Sites contributed a median of 1,167 patients with sepsis-associated MODS (interquartile range [IQR] 837, 1596). The median unadjusted site-specific in-hospital mortality was 9.9% (IQR 8.2%, 13.1%), which was significantly different across sites (p<0.001). Table E1 presents the clinical characteristics and outcomes of patients with and without sepsis-associated MODS in the cohort. Children with sepsis-associated MODS were randomly split by site into a derivation set (7 sites with 7,503 patients), a first external validation set (3 sites with 3,484 patients), and a second external validation set (3 sites with 4,259 patients).

### Derivation of trajectory-based candidate phenotypes using SANMF

Subgraph mining uncovered 776 frequent subgraphs representing individual organ dysfunction trajectories in patients with sepsis-associated MODS in the derivation set. 7,448 patients (99.3% of the derivation set) had at least one frequent subgraph and were included in the SANMF analysis. In the SANMF analysis, patients were separated into four trajectory-based groups based on the cophenetic correlation and the group size. Groups 1, 3, and 4 had an overall trajectory towards recovery of their organ dysfunctions in the first 72 hours (Figure E1) and associated in-hospital mortalities in the 4.3%-6.3% range. Group 2 represented the only high-risk group with an in-hospital mortality of 20.9% and was characterized by a trajectory of persistent organ dysfunction in the first 72 hours, predominantly respiratory, neurologic, and cardiovascular dysfunctions (Table E2).

### Reproducibility of the candidate phenotypes

Reproducibility of the SANMF groups in an external validation set was performed by applying the SANMF mixture coefficient matrix from the derivation set to the frequent subgraph counts of the first validation set. This demonstrated very reproducible distributions (i.e. proportion of patients in each group) and associated clinical outcome across the derivation and first validation sets (Table E2).

We then compared the overlap of the four SANMF-derived groups with the trajectory groups derived using RM-LCA and GBTM. Only SANMF Group 2 had significant overlap with the groups derived by RM-LCA and GBTM (Table E3). Specifically, RM-LCA Class 3 had a 78% overlap with SANMF Group 2 and GBTM Group 4 had 79% overlap with SANMF Group 2. The agreement between the models for these three groups was moderate (Fleiss’ Kappa = 0.5, p<0.001). All other overlaps ranged from 1 to 48% and had poor agreement.

### Clinical characterization and external reproducibility using a classifier

SANMF Group 2 was selected for further characterization as a sepsis phenotype because of its association with poor outcomes and its reproducibility across an external validation set and after using three different unsupervised trajectory modeling approaches. Based on the type, severity, and trajectory of the organ dysfunctions associated with the phenotype, we labeled it *persistent hypoxemia and shock* and grouped the rest of the patients under the label *other sepsis-associated MODS* (Figure 1).

**Figure 1.**
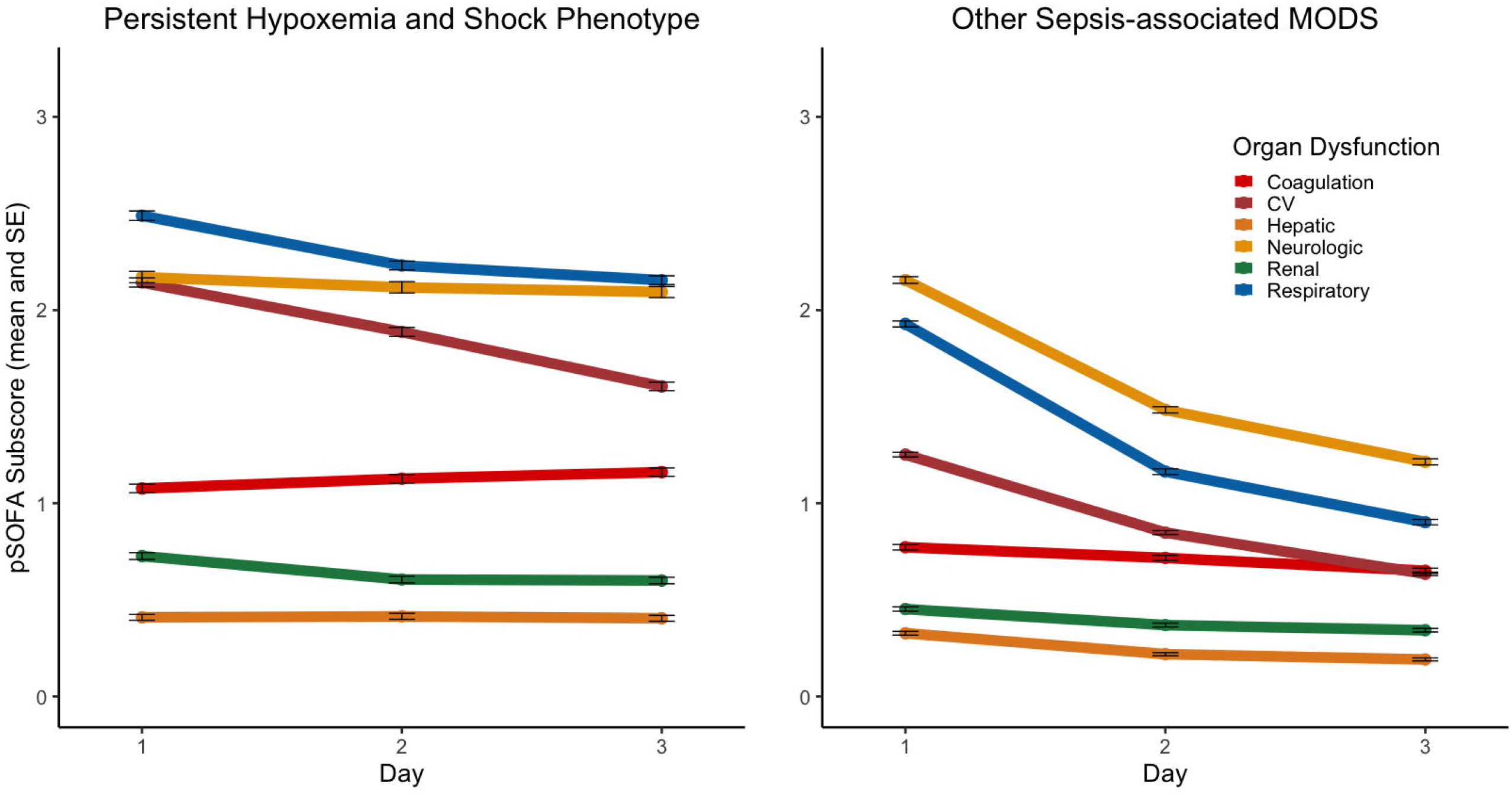
Type, severity, and trajectory of the organ dysfunctions associated with the *persistent hypoxemia and shock* phenotype.

We then trained a Random Forest classifier on the derivation set using the 18 daily pSOFA subscores as features (6 subscores per day for 3 days) and the *persistent hypoxemia and shock* phenotype as the outcome. The classifier had excellent performance at predicting the *persistent hypoxemia and shock* phenotype in the first validation set (area under the curve = 0.97, 95% confidence interval [CI] 0.96-0.97). The variable importance for the Random Forest is presented in Figure E2. When the model was applied to the second external validation set, patients classified as *persistent hypoxemia and shock* had a similar distribution, severity of illness, and outcomes when compared to patients with the phenotype in the derivation set and first validation set, with some differences in the rate of comorbidities and organ support use reflective of the baseline populations in the different sites (Table E4). Table 1 presents the clinical characteristics and outcomes of patients with *persistent hypoxemia and shock* phenotype compared to patients with other sepsis-associated MODS in the entire cohort.

**Table 1.** Clinical characteristics of children with the *persistent hypoxemia and shock* phenotype and those with *other sepsis-associated MODS*.

| Variables | Persistent<br>Hypoxemia and<br>Shock (n=4,836) | Other Sepsis-associated MODS<br>(n=10,410) |  |
| --- | --- | --- | --- |
| Age, years (IQR) | 4.3 (1, 11.7) | 5.2 (1.4, 11.8) | <0.001 |
| Male, No. (%) | 2,637 (54.5) | 5,742 (55.2) | 0.478 |
| Race/Ethnicity, No. (%) |  |  |  |
| White, Non-Hispanic | 2,279 (47.1) | 5,127 (49.3) | 0.009 |
| Black | 870 (18.0) | 1,754 (16.8) |  |
| Hispanic | 875 (18.1) | 1,966 (18.9) |  |
| Asian | 213 (4.4) | 424 (4.1) |  |
| Other | 599 (12.4) | 1,139 (10.9) |  |
| Comorbidities, No. (%) |  |  |  |
| Immunocompromised | 953 (19.7) | 1,955 (18.8) | 0.183 |
| Technology Dependent | 2,166 (44.8) | 4,733 (45.5) | 0.445 |
| Admission Source, No. (%) |  |  |  |
| Emergency Department | 2,000 (41.4) | 4,789 (46.0) | <0.001 |
| Hospital Floor | 1,091 (22.6) | 2,305 (22.1) |  |
| Direct/Transport | 1,307 (27.0) | 2,118 (20.3) |  |
| Operating Room | 438 (9.1) | 1,198 (11.5) |  |
| Season, No. (%) |  |  |  |
| Spring | 1,254 (25.9) | 2,686 (25.8) | 0.066 |
| Summer | 1,028 (21.3) | 2,262 (21.7) |  |
| Fall | 1,063 (22.0) | 2,442 (23.5) |  |
| Winter | 1,491 (30.8) | 3,020 (29.0) |  |
| Year, No. (%) |  |  |  |
| 2012-2013 | 1,466 (30.3) | 3,117 (29.9) | 0.563 |
| 2014-2015 | 1,601 (33.1) | 3,538 (34.0) |  |
| 2016-2017 | 1,769 (36.6) | 3,755 (36.1) |  |
| PRISM III score (IQR) | 15 (10, 23) | 9 (5, 14) | <0.001 |
| Organ support, No (%) |  |  |  |
| Mechanical ventilation | 3,787 (78.3) | 5,940 (57.1) | <0.001 |
| Vasoactive infusion | 3,348 (69.2) | 2,844 (27.3) | <0.001 |
| CRRT | 390 (8.1) | 208 (2.0) | <0.001 |
| ECMO | 337 (7.0) | 70 (0.7) | <0.001 |
| Outcomes |  |  |  |
| Length of stay, days (IQR) | 15 [7, 29] | 9 (5, 18) | <0.001 |
| Persistent MODS on day 7, No. (%) | 2,446 (50.6) | 1,722 (16.5) | <0.001 |
| In-hospital mortality, No. (%) | 1,045 (21.6) | 492 (4.7) | <0.001 |

Abbreviations: *IQR*, interquartile range; *PRISM III*, Pediatric risk of mortality version 3; *CRRT*, continuous renal replacement therapy; *ECMO*, extracorporeal membrane oxygenation.

### Prognostic relevance

In the mixed effects model, after adjusting for age, immunocompromised state, and mean pSOFA score in the first 72 hours, the *persistent hypoxemia and shock* phenotype was associated with a four-fold higher odds of in-patient mortality and a two-fold higher odds of persistent MODS at 7 days (Table 2). Figure E3 presents the Kaplan-Meier survival curve of the *persistent hypoxemia and shock* phenotype compared to patients with other sepsis-associated MODS.

**Table 2.** Prognostic relevance associated with the persistent hypoxemia and shock phenotype based on the generalized linear mixed effects models.

| Fixed effects | Adjusted OR* (95% CI) for In-Hospital Mortality | Adjusted OR* (95% CI) for Persistent MODS on day 7 |
| --- | --- | --- |
| Persistent hypoxemia and shock phenotype | 4 (2.9, 5.5) | 2 (1.6, 2.6) |
| Mean pSOFA by 72 hours | 1.4 (1.4, 1.5) | 1.8 (1.7, 1.9) |
| Age, years | 1 (0.9, 1) | 1 (1, 1) |
| Immunocompromised | 1.7 (1.2, 2.5) | 1 (0.7, 1.4) |

\*The random effect was the study site
Abbreviations: *OR*, odds ratio; *CI*, confidence interval; *MODS*, multiple organ dysfunction621 syndrome; *pSOFA*, pediatric sequential organ failure assessment.

### Therapeutic relevance

Overall, 14,983 patients survived for >24 hours and were included in the propensity matched analysis. The risk factors associated with higher likelihood of receiving hydrocortisone or albumin were: age, immunocompromised status, admission source, PRISM III score, vasoactive-inotropic score (VIS), and study site (Tables E5-E7). We used those factors to perform the propensity score matching (PSM) analysis. Using PSM, we matched 1,648 patients who received hydrocortisone (95%), and 1,162 patients who received albumin (100%) to untreated controls with similar propensity scores. Adequate covariate balance was achieved in both the hydrocortisone (Figure E4) and albumin (Figure E6) analyses. We found a significant interaction between the *persistent hypoxemia and shock* phenotype and both hydrocortisone and albumin use: treated patients with the phenotype had lower mortality and less MODS at 7 days compared to patients with other sepsis-associated MODS (Table 3). The Kaplan-Meier curves of treated and matched untreated patients with or without the *persistent hypoxemia and shock* phenotype are presented in Figures E5 and E7.

**Table 3.**
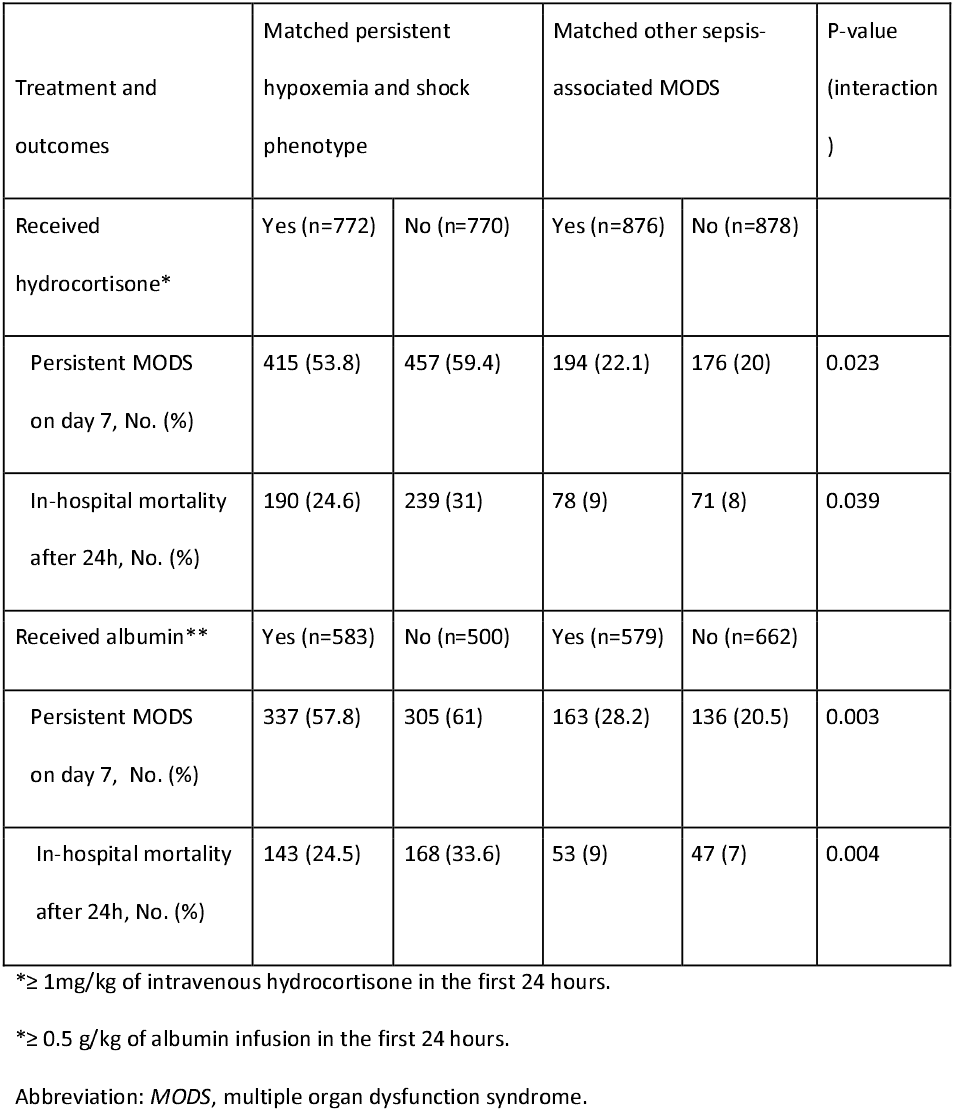
Therapeutic relevance associated with the persistent hypoxemia and shock phenotype based on the propensity score matched analysis.

In the sensitivity analysis using IPTW in the entire cohort with the same risk factors as in the PSM analysis, the interaction between the *persistent hypoxemia and shock* phenotype and hydrocortisone or albumin use were associated with lower in-hospital mortality (p<0.001).

### Pathobiological relevance

Patients with the *persistent hypoxemia and shock* had more systemic inflammation, coagulopathy, and signs of hypoperfusion than patients with other sepsis-associated MODS (Table 4). Among the 689 patients (4.5%) who had a ferritin level obtained, the majority of patients with *persistent hypoxemia and shock* had hyperferritinemia, with more than half reaching levels >1000 ng/mL (Table 4).

**Table 4.** Biochemical characteristics of the *persistent hypoxemia and shock phenotype* compared to other sepsis-associated MODS.

| Variables | Patients<br>with<br>values | Persistent<br>Hypoxemia and<br>Shock (n=4,836) | Other Sepsis-associated<br>MODS<br>(n=10,410) | p-value |
| --- | --- | --- | --- | --- |
| Laboratory results, median<br>(IQR)* |  |  |  |  |
| Min. Abs. lymphocytes, K/uL | 11,877 | 0.99 (0.43, 1.81) | 1.17 (0.54, 2.14) | <0.001 |
| Min. Hemoglobin, g/dL | 13,292 | 8.3 (7.1, 9.7) | 9.0 (7.5, 10.5) | <0.001 |
| Max. WBC, K/uL | 12,812 | 12.9 (7.8, 20.2) | 11.20 (6.9, 17) | <0.001 |
| Max. Bands, % | 6,205 | 13 (5, 25) | 9 (3, 19) | <0.001 |
| Max. CRP, mg/dL | 3,886 | 9.5 (3.3, 22.9) | 6.7 (2.5, 18.3) | <0.001 |
| Max. Ferritin, ng/mL | 689 | 1102 (315, 9407) | 435 (154, 2774) | <0.001 |
| Min. Platelets, K/uL | 12,768 | 105 (42, 199) | 149 (63, 238) | <0.001 |
| Max. INR | 7,713 | 1.6 (1.3, 2.1) | 1.3 (1.2, 1.7) | <0.001 |
| Max. PTT, sec. | 7,644 | 42 (33, 68) | 36 (30, 47) | <0.001 |
| Max. ALT, U/L | 8,722 | 53 (27, 161) | 40 (23, 91) | <0.001 |
| Max. AST, U/L | 8,639 | 86 (43, 308) | 54 (32, 128) | <0.001 |
| Max. Total Bilirubin, mg/dL | 8,669 | 0.8 (0.3, 2.3) | 0.6 (0.3, 2.1) | <0.001 |
| Min. Albumin, g/dL | 10,067 | 2.4 (2.0, 2.8) | 2.6 (2.2, 3.1) | <0.001 |
| Max. BUN, g/dL | 14,407 | 15 (9, 26) | 11 (7, 18) | <0.001 |
| Max. Creatinine, g/dL | 14,267 | 0.5 (0.3, 0.9) | 0.4 (0.3, 0.6) | <0.001 |
| Max. Glucose, g/dL | 14,225 | 196 (143, 285) | 148 (117, 206) | <0.001 |
| Min. pH | 12,037 | 7.21 (7.10, 7.29) | 7.30 (7.23, 7.35) | <0.001 |
| Min. Bicarbonate, mEq/L | 14,584 | 19 (15, 22) | 21 (18, 23) | <0.001 |
| Max. Lactate, mmol/L | 9,294 | 2.9 (1.6, 6.3) | 1.8 (1.1, 3.1) | <0.001 |
| Min. PaO<sub>2</sub>, mmHg | 6,726 | 63 (52, 76) | 74 (59, 95) | <0.001 |
| Max. PaCO<sub>2</sub>, mmHg | 6,726 | 55 (45, 68) | 46 (40, 54) | <0.001 |

\*minimum (min.) or maximum (max.) value within 72 hours of admission, as indicated
Abbreviations: *IQR*, inter-quartile range; *Abs.*, absolute; *WBC*, white blood cell count; *CRP*, c-reactive protein ; *INR*, international normalized ratio; *PTT*, partial thromboplastin time ; *ALT*, alanine aminotransferase; *AST*, aspartate aminotransferase ; *BUN*, blood urea nitrogen; *PaO<sub>2</sub>*, partial pressure of arterial oxygen; *PaCO<sub>2</sub>*, partial pressure of arterial carbon dioxide.

When compared to other high-risk clinical syndromes in the first 72 hours of admission, the *persistent hypoxemia and shock* phenotype overlapped with 55% of patients meeting septic shock criteria, 54% of patients meeting moderate-to-severe pediatric ARDS criteria, and 62% of patient in the top tertile of organ dysfunction burden based on the mean pSOFA score (Table E8).

Finally, the *persistent hypoxemia and shock* phenotype was more common in younger patients, especially <1 year old, where the prevalence was 36-45% compared to 28-32% in the older age groups (Table E9).

## DISCUSSION

In this study, we derived and validated the *persistent hypoxemia and shock* phenotype in children with sepsis-associated MODS. This trajectory-based sepsis phenotype had features of systemic inflammation and coagulopathy, was present in approximately one third of children with sepsis-associated MODS, and was independently associated with mortality. Classification of this phenotype was highly reproducible in two external validation sets and using various statistical methods for trajectory modeling. Additionally, in a propensity score analysis, patients with *persistent hypoxemia and shock* appeared to have a higher likelihood to benefit from adjuvant therapy with hydrocortisone and albumin when compared to other patients with sepsis-associated MODS. Finally, the *persistent hypoxemia and shock* phenotype overlapped with only about half of the patients with septic shock, half of those with moderate-to-severe pediatric ARDS, and less than two thirds of those with the highest organ dysfunction burden based on the mean pSOFA score in the first 72 hours. These latter findings highlight the fact that this phenotype does not simply describe the sickest patients or those recognizable as septic shock or ARDS at the bedside, instead it suggests that the *persistent hypoxemia and shock* phenotype represents a distinct clinical phenotype of sepsis-associated MODS with a disproportionately high risk of mortality, accounting for almost 70% of all deaths in patients in this cohort.

Prior studies have identified phenotypes with characteristics similar to the *persistent hypoxemia and shock* phenotype. Knox et al. described the *shock with hypoxemia and altered mental status* phenotype and Seymour et al. described the γ phenotype in adult sepsis patients, which share similar features, including hypoxemia, vasoactive dependence, inflammation, and high mortality [12,13]. In follow up studies, the γ phenotype has been found to be particularly predominant in patients with viral and bacterial pneumonia, which are common etiologies of sepsis and respiratory failure in children [29]. Calfee et al. and Dahmer et al. have also described a *hyperinflammatory* phenotype of ARDS in adults and children, respectively, which share many common features with the *persistent hypoxia and shock* phenotype [30,31]. Similar to our findings, the *hyperinflammatory* ARDS phenotype is commonly associated with more sepsis, inflammation, hypotension, and coagulopathy than other patients with ARDS [30–32].

Importantly, Calfee and colleagues have shown that the *hyperinflammatory* ARDS phenotype is associated with heterogeneity of treatment effect to various common interventions, including ventilator strategies and fluid management [30,33,34]. Additionally, our group previously described a high-risk phenotype with severe hypoxemia and shock in children with MODS (both with and without sepsis) in a two-center study [3]. On a related concept, Villar et al. performed an enrichment strategy in their ARDS randomized controlled trial by only enrolling adult patients who had persistent hypoxemia 24 hours after initial ARDS diagnosis and found that patients who met that persistent hypoxemia criteria and received dexamethasone had a significantly lower mortality than controls [35]. Finally, Carcillo et al. have described the hyperferritinemic response in sepsis as a distinct, high-risk phenotype characterized by macrophage activation that is potentially susceptible to anti-cytokine therapy [8,36,37]. Importantly, macrophage activation appears to be a key mechanism in sepsis-related lung injury [38–41].

Our findings have important implications. While risk stratifying critically ill children with infections and sepsis early in the course is important for diagnostic purposes, the clinical reality is that most children with sepsis will suffer their worst degree of MODS on the day of admission. Most of them, even those with significant organ dysfunction, will tend to follow a trajectory of recovery after admission, as observed in our cohort and in previous studies [4,42]. Thus, understanding which children with sepsis will have a trajectory of persistent or worsening organ dysfunction, what patterns of dysfunction are expected in those children, and why they display those patterns from a pathobiological standpoint, become questions of paramount importance. In our study we have identified a high-risk, highly reproducible trajectory-based phenotype of pediatric sepsis with both prognostic and therapeutic relevance. However, the *persistent hypoxemia and shock* phenotype requires longitudinal information for patient classification. In order for this type of classification to be clinically useful it would need to be predicted earlier in the clinical course.

Prediction of phenotype membership is the next, important step in this line of research and could be performed through a combination of machine learning approaches using EHR data [43], rapid-turnaround biomarkers (including ferritin and novel cytokine profiles) [44], and other physiological markers like low heart rate variability, which is associated with pro-inflammatory states and progressive organ dysfunction [45,46].

Our study has several strengths and limitations. We performed our analysis using a large, granular, multicenter cohort with wide geographic and racial-ethnic representation of children in the United States. Furthermore, we derived and validated the *persistent hypoxemia and shock* phenotypes using data that was partitioned by study sites, which allowed us to test the external validity and generalizability of the phenotype. However, our dataset was observational in nature, and thus susceptible to selection bias and the uncertainty introduced by missing data. While we used standard approaches to ensure data quality, deal with missing data, and adjust for confounders, further validation of our findings is needed. Finally, our therapeutic relevance analysis using propensity scoring is susceptible to residual confounding and must be interpreted with caution.

In conclusion, we derived and validated the *persistent hypoxemia and shock* phenotype, a trajectory-based organ dysfunction phenotype which was present in a third of children with sepsis-associated MODS and associated with more than two thirds of deaths. This phenotype is associated with systemic inflammation and coagulopathy, is independently associated with higher mortality, and is associated with heterogeneity of treatment effect to common adjuvant therapies. Future studies are needed to ascertain the reproducibility of this phenotype, assess whether it can be predicted earlier in the course, further study the possible biological mechanisms underlying it, and investigate candidate therapeutic targets.

## Data Availability

All data produced in the present study are available upon reasonable request to the authors

## ACKNOWLEDGEMENTS

The authors would like to acknowledge and thank the late Dr. Hector Wong (1963-2022), who was an early collaborator and supporter of this work.

## AUTHOR CONTIRBUTIONS

*Concept, design and drafting of manuscript*: LNS; *Analysis*: LNS, ES; *Acquisition and/or interpretation of the data, revising manuscript critically for important intellectual content and final approval of the version to be published*: All authors

## AVAILABILITY OF DATA

The datasets generated and analyzed during the current study are not publicly available due institutional review board restrictions given that it contains personal health information, but are available from the corresponding author on reasonable request.

## DECLARATIONS

- The institutional review board at Ann & Robert H. Lurie Children’s Hospital of Chicago served as the central review board for this study and approved it with a waiver of consent.
- The authors declare no financial conflicts of interest related to this study.

