## Supplemental Information for "Derivation, validation, and clinical relevance of a pediatric sepsis phenotype with persistent hypoxemia and shock"

**Content**

Page 2. **Additional file - Methods**

Page 4. **Additional file - Table 1**. Clinical characteristics of critically ill children with confirmed or suspected infection with or without sepsis-associated multiple organ dysfunction syndrome (MODS).

Page 6. **Additional file - Figure 1**. Alluvial plots of the SANMF groups based on average pSOFA subscores.

Page 7. **Additional file - Table 2.** Distribution and clinical outcomes of the SANMF groups in the derivation and first validation set.

Page 8. **Additional file - Table 3**. Overlap of RM-LCA classes and GBTM groups with SANMF groups amongst patients in the derivation and first validation set.

Page 9. **Additional file - Figure 2**. Variable importance plot for the Random Forest classifier.

Page 10. **Additional file - Table 4**. Clinical characteristics, comorbidities, organ support, and outcomes of patients with the *persistent hypoxemia and shock* phenotype in the derivation and validation sets.

Page 11. **Additional file - Figure 3**. Kaplan-Meier survival curve for patients with the *persistent hypoxemia and shock* phenotype and those with *other sepsis-associated-MODS.*

Page 12. **Additional file - Table 5**. Univariate analysis of factors associated with hydrocortisone administration in patients who were alive for the first 24 hours and were included in the therapeutic relevance analysis.

Page 13. **Additional file - Table 6**. Univariate analysis of factors associated with albumin administration in patients who were alive for the first 24 hours and were included in the therapeutic relevance analysis.

Page 14. **Additional file - Table 7.** Univariate analysis of factors associated with death in patients who were alive for the first 24 hours and were included in the therapeutic relevance analysis.

Page 15. **Additional file - Figure 4**. Covariate balance before and after propensity score matching in patients in the hydrocortisone analysis.

Page 16. **Additional file - Figure 5**. Kaplan-Meier survival curve for patients with the *persistent hypoxemia and shock* phenotype and those with *other sepsis-associated-MODS* who were treated or not with hydrocortisone.

Page 17. **Additional file - Figure 6**. Covariate balance before and after propensity score matching in patients in the albumin analysis.

Page 18. **Additional file - Figure 7**. Kaplan-Meier survival curve for patients with the *persistent hypoxemia and shock* phenotype and with *other sepsis-associated-MODS* treated or not with albumin.

Page 19. **Additional file - Table 8**. Distribution of patients with vasoactive-dependent shock, moderate-to-severe pediatric ARDS, and in top tertile of the mean pSOFA score across the *persistent hypoxemia and shock* phenotype and patients with other sepsis-associated MODS in the first 72 hours of admission.

Page 20. **Additional file - Table 9**. Distribution of patients with the *persistent hypoxemia and shock* phenotype across age groups.

Page 21. **References**.

**Additional file - Methods**

***Subgraph-Augmented Nonnegative Matrix Factorization (SANMF)***

*Overview*. Nonnegative matrix factorization (NMF) is an algorithm with clustering properties that is designed to explain the observed data using a limited number of hidden features that when combined together approximate the original data as accurately as possible (1). NMF-derived features are simple, additive, and intuitive, which greatly increases their interpretability. SANMF enhances NMF by incorporating subgraph mining in order to group similar trajectories in the data, in our case the trajectories of the six pSOFA subscores during the first 3 days of critical illness in patients with sepsis-associated MODS. The methodological approach was based on previous analyses by our group (2, 3).

*Subgraph Mining*. Node-edge list graphs were prepared for patients with sepsis-associated MODS by mapping each pSOFA subscore on days 1, 2, and 3 as nodes, and the edges as the transitions between days 1-to-2 and 2-to-3. Edges were labeled with the direction of change between subsequent nodes, and were not day-sensitive. Common subgraphs were extracted from the node-edge list graphs generated using the Molecular Substructure Miner (MoSS) developed by Borgelt et al. (4). The MoSS search algorithm was run with no complement group and with no frequency requirement to mine as many potential subgraphs as possible. The number of subgraphs was then reduced by eliminating: (1) small subgraphs contained within larger subgraphs in order to reduce double-counting; (2) subgraphs that were static and did not have a maximum subscore of more than 2 in order to reduce the influence of organs with mild or no dysfunction and no change; and (3) subgraphs present in less than 20 patients in order to focus on more representative subgraphs. Patients who had no subgraphs remaining after the above step were eliminated from the remainder of the analysis.

*Non-negative Matrix Factorization*. NMF was implemented on the derivation set enforcing sparsity using the SNMF/L algorithm with non-negative double singular value decomposition seeding as previously described (1, 2). The total number of phenotypes was determined using a combination of cophenetic correlation and a pragmatic approach based on group size with a minimal group size of 10% of the derivation cohort required. This resulted in a matrix of hidden features, used to define the candidate phenotypes, and a mixture coefficient matrix that was used to assess the reproducibility of the candidate phenotypes in the first validation set, as further described in the main text.

***Repeated Measures Latent Class Analysis (RM-LCA)***

RM-LCA has been used as a patient-centered approach to identify patterns of clinical or behavioral change in a time-dependent manner (5, 6). The 18 daily pSOFA subscores were used as features and the polytomous latent class analysis algorithm of the *poLCA* R package were used to derive the latent classes, with ten random starting values and 1,000 maximum iterations of the estimation algorithm (7). Four classes were chosen based on the results of the SANMF analysis.

***Group-based Trajectory Modeling (GBTM)***

GBTM has been used to identify naturally occurring trajectory groups based on one or more variables, including in multiple critical care studies (8–10). Data were analyzed using the *traj* plugin for GBTM analysis in STATA. We fitted a model using four groups with the censored normal distribution and cubic polynomials based on the Bayesian information criterion (BIC) after assessing the fit of linear, quadratic, cubic, and quartic polynomials.

***Random Forest (RF) Classifier***

An RF classifier was trained to predict phenotype membership using the 18 daily pSOFA subscores. The *caret* R package was used to train the RF classifier using 10-fold cross-validation and 1,000 trees. The *ggplot2* R package was used to plot the variable importance of the mode.

***Propensity Score Matched Analysis***

Propensity score matching (PSM) is a quasi-experimental method in which a control group that is comparable to the treatment group is developed by matching each treated patient with a non-treated patient of similar characteristics based on a propensity score. For the response to adjuvant therapy analysis using propensity score matching, we studied intravenous hydrocortisone (≥ 1mg/kg in the first 24 hours) and albumin (≥ 0.5 g/kg in the first 24 hours). We used the *matchIt* R package to perform the PSM using 1:1 nearest neighbor matching based on a caliper of 0.2 standard deviations of the logit of the propensity score. The propensity score to receive either of the studied treatments in the first 24 hours of admission was based on risk factors that were significantly associated with a higher likelihood of receiving the treatment with a p<0.05 in univariate analysis in the patients included in the analysis. To limit the influence of survival bias (i.e. where patients who receive the treatment appear to have improved survival simply because untreated patients did not survive long enough to receive the therapy), only patients who were alive for at least 24 hours were included in the PSM. After the matching, differences in outcomes (in-hospital mortality and persistent MODS on day 7) based on phenotype membership in treated and untreated patients were assessed using logistic regression models with an interaction term for treatment and phenotype.

***Inverse Probability Treatment Weighting (IPTW)***

As a sensitivity analysis of the response to adjuvant therapy analysis using PSM using all patients, we performed an analysis using IPTW using the same variables as in the PSM. While PSM is a commonly used technique to adjust for selection bias, the process of matching untreated patients as controls can result in significant information loss given that many untreated patients will remain unmatched. To avoid this potential information loss, we performed IPTW using the *ipw* R package to weigh every eligible patient in the treatment analysis by their inverse probability of receiving the treatment using the same confounders as in the PSM analysis using logistic regression. Again to avoid survival bias, only patients alive for at least 24 hour were included in the analysis. After a weight was assigned to each patient, we used the *survey* R package to perform weighted interaction analyses between treatment and phenotype to assess the differences in outcomes in treated and untreated patients by adjusting for selection bias with the IPTW-based weights.

**Additional file - Table 1**. Clinical characteristics of critically ill children with confirmed or suspected infection with or without sepsis-associated multiple organ dysfunction syndrome (MODS).

| Variables | Sepsis-associated MODS | | p-value |
| --- | --- | --- | --- |
|  | Yes (n=15,246) | No (n=23,486) |  |
| Age, years (IQR) | 4.9 (1.3, 11.8) | 4.2 (1.2, 10.8) | <0.001 |
| Male, No. (%) | 8,379 (55.0) | 12,827 (54.6) | 0.51 |
| Race/Ethnicity, No. (%) |  |  |  |
| White, Non-Hispanic | 7,406 (48.6) | 11,497 (49.0) | <0.001 |
| Black | 2,624 (17.2) | 4,557 (19.4) |  |
| Hispanic | 2,841 (18.6) | 4,170 (17.8) |  |
| Asian | 637 (4.2) | 1,052 (4.5) |  |
| Other | 1,738 (11.4) | 2,210 (9.4) |  |
| Comorbidities, No. (%) |  |  |  |
| Immunocompromised | 2,908 (19.1) | 3,189 (13.6) | <0.001 |
| Technology Dependent | 6,899 (45.3) | 8,174 (34.8) | <0.001 |
| Admission Source, No. (%) |  |  |  |
| Emergency Department | 6,789 (44.5) | 12,599 (53.6) | <0.001 |
| Hospital Floor | 3,396 (22.3) | 4,260 (18.1) |  |
| Direct/Transport | 3,425 (22.5) | 3,764 (16.0) |  |
| Operating Room | 1,636 (10.7) | 2,863 (12.2) |  |
| Season, No. (%) |  |  |  |
| Spring | 3,940 (25.8) | 6,092 (25.9) | 0.22 |
| Summer | 3,290 (21.6) | 5,125 (21.8) |  |
| Fall | 3,505 (23.0) | 5,538 (23.6) |  |
| Winter | 4,511 (29.6) | 6,731 (28.7) |  |
| Year, No. (%) |  |  |  |
| 2012-2013 | 4,583 (30.1) | 7,643 (32.5) | <0.001 |
| 2014-2015 | 5,139 (33.7) | 7,492 (31.9) |  |
| 2016-2017 | 5,524 (36.2) | 8,351 (35.6) |  |
| PRISM III score (IQR) | 11 (7, 17) | 4 (2, 7) | <0.001 |
| Length of stay, days (IQR) | 10.7 (5.5, 21.3) | 5.0 (2.8, 9.9) | <0.001 |
| In-hospital mortality, No. (%) | 1,537 (10.1) | 277 (1.2) | <0.001 |

Abbreviations: *IQR*, interquartile range; *PRISM III*, Pediatric risk of mortality version 3.

**Additional file - Figure 1**. Alluvial plots of the four SANMF groups* based on the average daily pSOFA subscores.

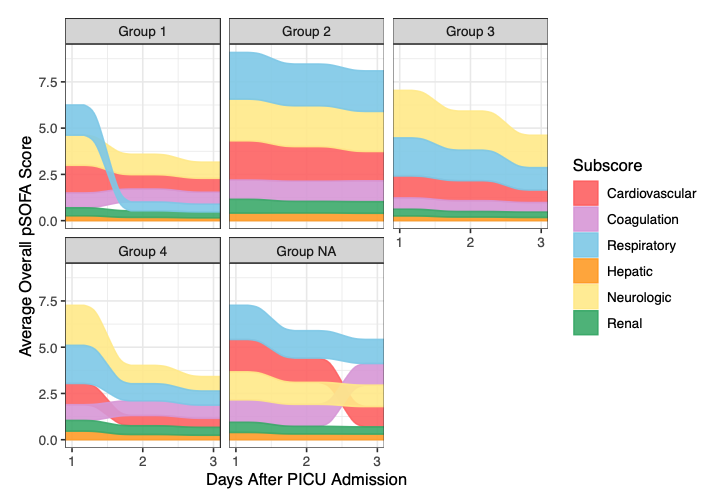

*The ‘Group 2’ represents the novel *persistent hypoxemia and shock* phenotype. The ‘Group NA’ plot represents the average daily pSOFA subscores for the 55 patients (0.7% of the derivation cohort) who had no frequent subgraphs and were excluded from the SANMF analysis.

Abbreviations: *SANMF*, subgraph augmented nonnegative matrix factorization; *pSOFA*, pediatric sequential organ failure assessment.

**Additional file - Table 2.** Distribution and clinical outcomes of the SANMF groups in the derivation and first validation set.

| SANMF Groups | Group 1 | | Group 2 | | Group 3 | | Group 4 | |
| --- | --- | --- | --- | --- | --- | --- | --- | --- |
|  | Derivation | Validation 1 | Derivation | Validation 1 | Derivation | Validation 1 | Derivation | Validation 1 |
| Total (% of set) | 1,050 (14.0) | 564 (16.2) | 2,529 (33.7) | 1,122 (32.2) | 2,134 (28.4) | 1,165 (33.4) | 1,735 (23.1) | 590 (16.9) |
| Length of stay,  median days (IQR) | 8 (5, 15) | 8 (5, 16) | 15 (7, 29) | 14 (7, 28) | 11 (6, 20) | 10 (6, 18) | 10 (5, 19) | 9 (4, 19) |
| Persistent MODS on day 7, No. (%) | 89 (8.5) | 57 (10.1) | 1,188 (47.0) | 584 (52.0) | 444 (20.8) | 226 (19.4) | 273 (15.7) | 99 (16.8) |
| In-hospital mortality, No. (%) | 45 (4.3) | 16 (2.8) | 528 (20.9) | 257 (22.9) | 134 (6.3) | 48 (4.1) | 92 (5.3) | 33 (5.6) |

Abbreviations: *SANMF*, subgraph augmented nonnegative matrix factorization; *IQR*, interquartile range; *MODS*, multiple organ dysfunction syndrome.

**Additional file - Table 3**. Overlap of RM-LCA classes and GBTM groups with SANMF groups amongst patients in the derivation and first validation set.

| SANMF  Groups | RM-LCA Classes | | | | GBTM Groups | | | |
| --- | --- | --- | --- | --- | --- | --- | --- | --- |
|  | Class 1 (n=2,241) | Class 2 (n=2,891) | Class 3 (n=1,746) | Class 4 (n=4,011) | Group 1 (n=2,241) | Group 2 (n= 6,005) | Group 3 (n=1,057) | Group 4 (n=1,586) |
| Group 1 | 467 (21*) | 644 (22) | 18 (1) | 485 (12) | 498 (22) | 955 (16) | 142 (13) | 19 (1) |
| Group 2 | 630 (28) | 591 (20) | **1,367 (78)** | 1,063 (27) | 598 (27) | 1,448 (24) | 358 (34) | **1,247 (79)** |
| Group 3 | 503 (22) | 604 (21) | 249 (14) | 1,943 (48) | 545 (24) | 2,272 (38) | 247 (23) | 235 (15) |
| Group 4 | 641 (29) | 1,052 (36) | 112 (6) | 520 (13) | 600 (27) | 1,330 (22) | 310 (29) | 85 (5) |

*Percentage (%) overlap of RM-LCA or GBTM group patients with those in the SANMF groups.

Abbreviations: *RM-LCA*, repeated measures latent class analysis; *GBTM*, group-based trajectory modeli

**Additional file - Figure 2**. Variable importance plot for the Random Forest classifier.

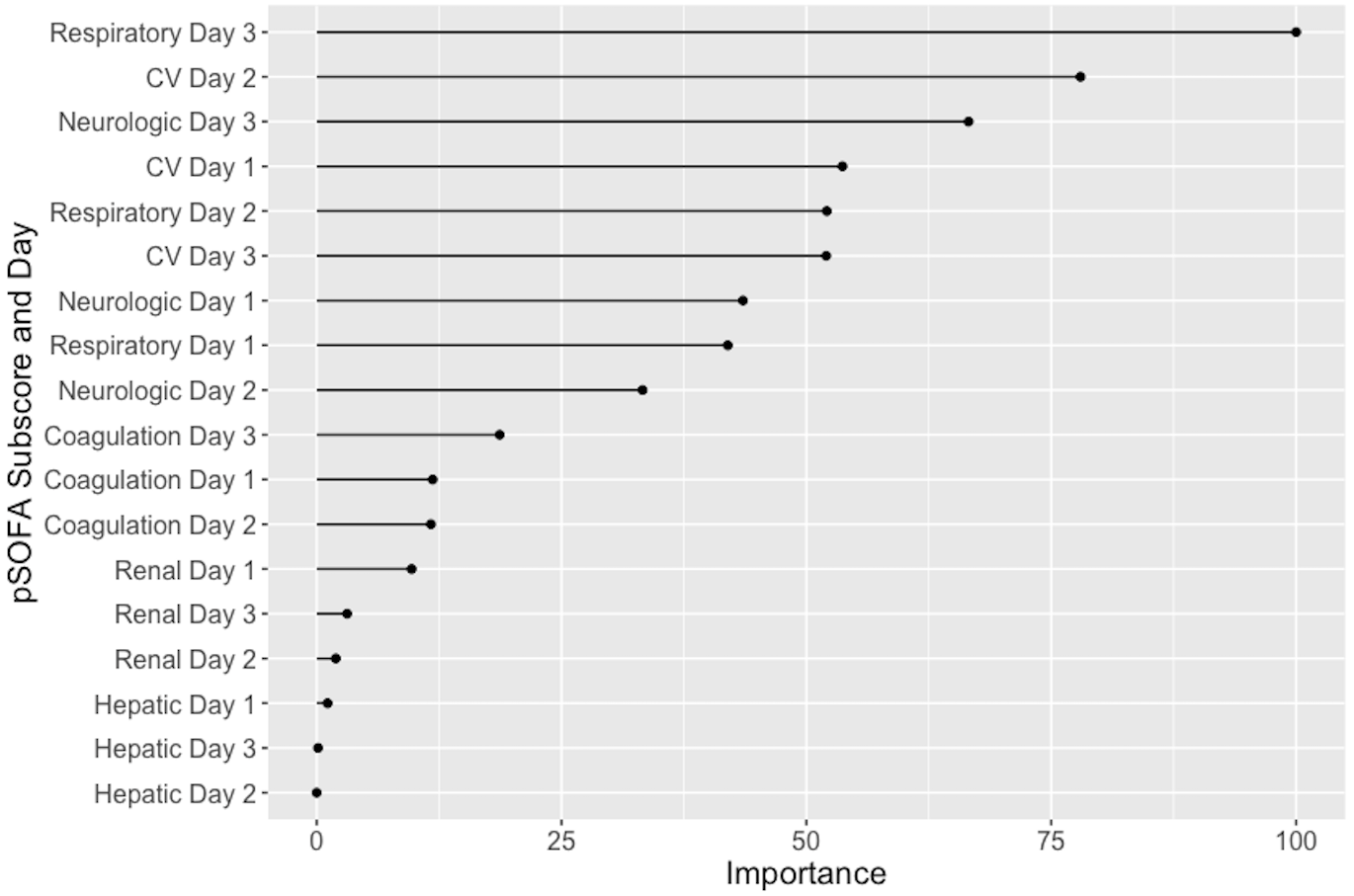

Abbreviations: *pSOFA*, pediatric sequential organ failure assessment; *CV*, cardiovascular.

**Additional file - Table 4**. Clinical characteristics, comorbidities, organ support, and outcomes of patients with the *persistent hypoxemia and shock* phenotype in the derivation set and two validation sets.

| Variables | Derivation set (n=7,503) | First validation set (n=3,484) | Second validation set (n=4,259) |
| --- | --- | --- | --- |
| Total No. (% of set) | 2,529 (33.7) | 1,122 (32.2) | 1,185 (27.8) |
| Age, year (IQR) | 5.2 (1.3, 12.3) | 3 (0.5, 10.6) | 3.6 (0.8, 11.2) |
| PRISM III score (IQR) | 15 (9, 22) | 17 (11, 25) | 16 (10, 24) |
| Comorbidities |  |  |  |
| Immunocompromised (%) | 567 (22.4) | 170 (15.2) | 216 (18.2) |
| Technology Dependent (%) | 1085 (42.9) | 393 (35.0) | 688 (58.1) |
| Organ support, No. (%) |  |  |  |
| Mechanical ventilation | 2069 (81.8) | 659 (58.7) | 1059 (89.4) |
| Vasoactive infusion | 1733 (68.5) | 809 (72.1) | 806 (68.0) |
| ECMO | 192 (7.6) | 39 (3.5) | 106 (8.9) |
| CRRT | 231 (9.1) | 79 (7.0) | 80 (6.8) |
| Outcomes |  |  |  |
| Length of stay,  median days (IQR) | 15 (7, 29) | 14 (7, 28) | 15 (7, 29) |
| Persistent MODS on day 7, No. (%) | 1,188 (47.0) | 584 (52.0) | 674 (56.9) |
| In-hospital mortality, No. (%) | 528 (20.9) | 257 (22.9) | 260 (21.9) |

Abbreviations: *IQR*, interquartile range; *PRISM III*, Pediatric risk of mortality version 3; *ECMO*, extracorporeal membrane oxygenation; *CRRT*, continuous renal replacement therapy; *MODS*, multiple organ dysfunction syndrome.

**Additional file - Figure 3**. Kaplan-Meier survival curve for patients with the *persistent hypoxemia and shock* phenotype and those with *other sepsis-associated-MODS*

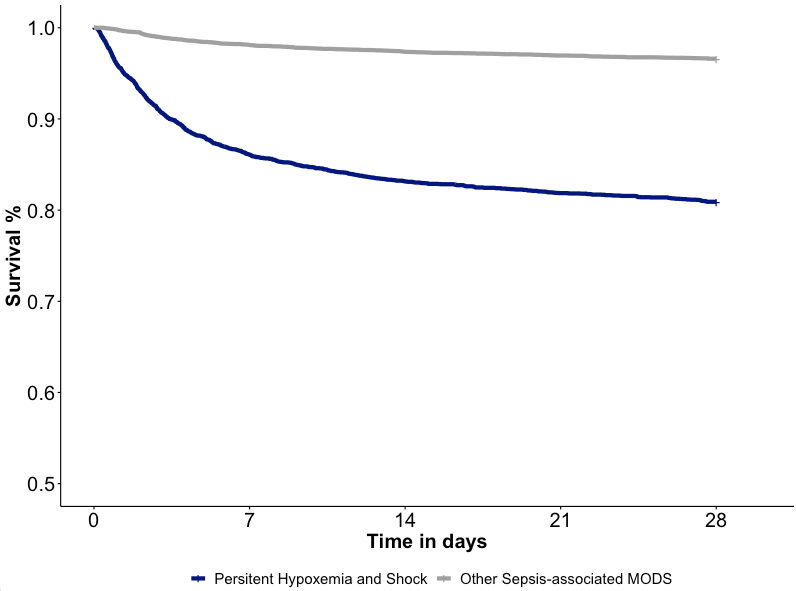

Abbreviations: *MODS*, multiple organ dysfunction syndrome.

**Additional file - Table 5**. Univariate analysis of factors associated with hydrocortisone administration in patients who were alive for the first 24 hours and were included in the therapeutic relevance analysis.

| Variables | Received hydrocortisone* | | p-value |
| --- | --- | --- | --- |
|  | Yes (n=1,734) | No (n=13,249) |  |
| Age, years (IQR) | 6 (2, 12) | 4 (1, 11) | <0.001 |
| Comorbidities, No. (%) |  |  |  |
| Immunocompromised | 509 (29.4) | 2,365 (17.9) | <0.001 |
| Technology Dependent | 848 (48.9) | 5,959 (45.0) | 0.002 |
| Admission Source, No. (%) |  |  |  |
| Emergency Department | 842 (48.6) | 5,826 (44.0) | <0.001 |
| Hospital Floor | 432 (24.9) | 2,927 (22.1) |  |
| Direct/Transport | 355 (20.5) | 2,983 (22.5) |  |
| Operating Room | 105 (6.1) | 1,513 (11.4) |  |
| Season, No. (%) |  |  |  |
| Spring | 412 (23.8) | 3,454 (26.1) | 0.123 |
| Summer | 401 (23.1) | 2,830 (21.4) |  |
| Fall | 410 (23.6) | 3,034 (22.9) |  |
| Winter | 511 (29.5) | 3,931 (29.7) |  |
| Year, No. (%) |  |  |  |
| 2012-2013 | 478 (27.6) | 4,011 (30.3) | 0.061 |
| 2014-2015 | 612 (35.3) | 4,434 (33.5) |  |
| 2016-2017 | 644 (37.1) | 4,804 (36.3) |  |
| PRISM III score (IQR) | 16 (10, 24) | 10 (6, 15) | <0.001 |
| Vasoactive-Inotropic Score, highest in first 24 hours (IQR) | 12 (0, 28) | 0 (0, 4) | <0.001 |

*≥ 1mg/kg of intravenous hydrocortisone in the first 24 hours.

Abbreviations: *IQR*, interquartile range; *PRISM III*, Pediatric risk of mortality version 3.

**Additional file - Table 6**. Univariate analysis of factors associated with albumin administration in patients who were alive for the first 24 hours and were included in the therapeutic relevance analysis.

| Variables | Received albumin* | | p-value |
| --- | --- | --- | --- |
|  | Yes (n=1,162) | No (n=13,821) |  |
| Age, years (IQR) | 4 (1, 10) | 5 (1, 11) | 0.032 |
| Comorbidities, No. (%) |  |  |  |
| Immunocompromised | 312 (26.9) | 2,562 (18.5) | <0.001 |
| Technology Dependent | 502 (43.2) | 6,305 (45.6) | 0.119 |
| Admission Source, No. (%) |  |  |  |
| Emergency Department | 446 (38.4) | 6,222 (45.0) | <0.001 |
| Hospital Floor | 290 (25.0) | 3,069 (22.2) |  |
| Direct/Transport | 241 (20.7) | 3,097 (22.4) |  |
| Operating Room | 185 (15.9) | 1,433 (10.4) |  |
| Season, No. (%) |  |  |  |
| Spring | 303 (26.1) | 3,563 (25.8) | 0.259 |
| Summer | 269 (23.1) | 2,962 (21.4) |  |
| Fall | 273 (23.5) | 3,171 (22.9) |  |
| Winter | 317 (27.3) | 4,125 (29.8) |  |
| Year, No. (%) |  |  |  |
| 2012-2013 | 378 (32.5) | 4,111 (29.7) | 0.034 |
| 2014-2015 | 354 (30.5) | 4,692 (33.9) |  |
| 2016-2017 | 430 (37.0) | 5,018 (36.3) |  |
| PRISM III score (IQR) | 17 (11, 24) | 10 (6, 16) | <0.001 |
| Vasoactive-Inotropic Score (IQR) | 8 (0, 21) | 0 (0, 5) | <0.001 |

*≥ 0.5 g/kg of intravenous albumin in the first 24 hours.

Abbreviations: *IQR*, interquartile range; *PRISM III*, Pediatric risk of mortality version 3.

**Additional file - Table 7.** Univariate analysis of factors associated with death in patients who were alive for the first 24 hours and were included in the therapeutic relevance analysis.

| Variables | Died in-hospital (after 24 hours) | | p-value |
| --- | --- | --- | --- |
|  | Yes (n=1,333) | No (n=13,650) |  |
| Age, years (IQR) | 4 (0, 11) | 4 (1, 11) | 0.029 |
| Comorbidities, No. (%) |  |  |  |
| Immunocompromised | 405 (30.4) | 2469 (18.1) | <0.001 |
| Technology Dependent | 581 (43.6) | 6226 (45.6) | 0.165 |
| Admission Source, No. (%) |  |  |  |
| Emergency Department | 458 (34.4) | 6210 (45.5) | <0.001 |
| Hospital Floor | 413 (31.0) | 2946 (21.6) |  |
| Direct/Transport | 389 (29.2) | 2949 (21.6) |  |
| Operating Room | 73 (5.5) | 1545 (11.3) |  |
| Season, No. (%) |  |  |  |
| Spring | 336 (25.2) | 3530 (25.9) | <0.001 |
| Summer | 313 (23.5) | 2918 (21.4) |  |
| Fall | 348 (26.1) | 3096 (22.7) |  |
| Winter | 336 (25.2) | 4106 (30.1) |  |
| Year, No. (%) |  |  |  |
| 2012-2013 | 393 (29.5) | 4096 (30.0) | 0.889 |
| 2014-2015 | 448 (33.6) | 4598 (33.7) |  |
| 2016-2017 | 492 (36.9) | 4956 (36.3) |  |
| PRISM III score (IQR) | 20 (13, 30) | 10 (6, 15) | <0.001 |
| Vasoactive-Inotropic Score (IQR) | 5 (0, 22) | 0 (0, 5) | <0.001 |

Abbreviations: *IQR*, interquartile range; *PRISM III*, Pediatric risk of mortality version 3.

**Additional file - Figure 4**. Covariate balance before and after propensity score matching in patients in the hydrocortisone analysis.

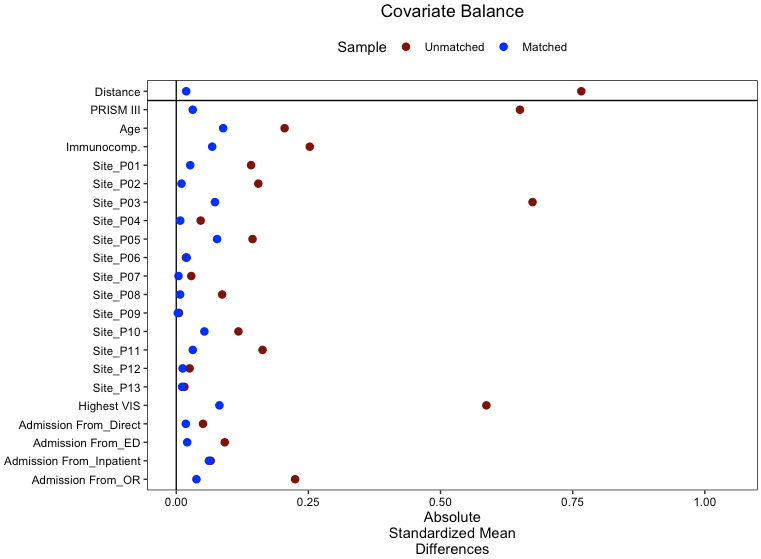

Abbreviations: *PRISM III*, Pediatric risk of mortality version 3; *Immunocomp.*, immunocompromised comorbidity.

**Additional file - Figure 5**. Kaplan-Meier survival curve* for patients with the *persistent hypoxemia and shock* (PHS) phenotype and those with *other sepsis-associated-MODS* (SA-MODS)who were treated or not with hydrocortisone.

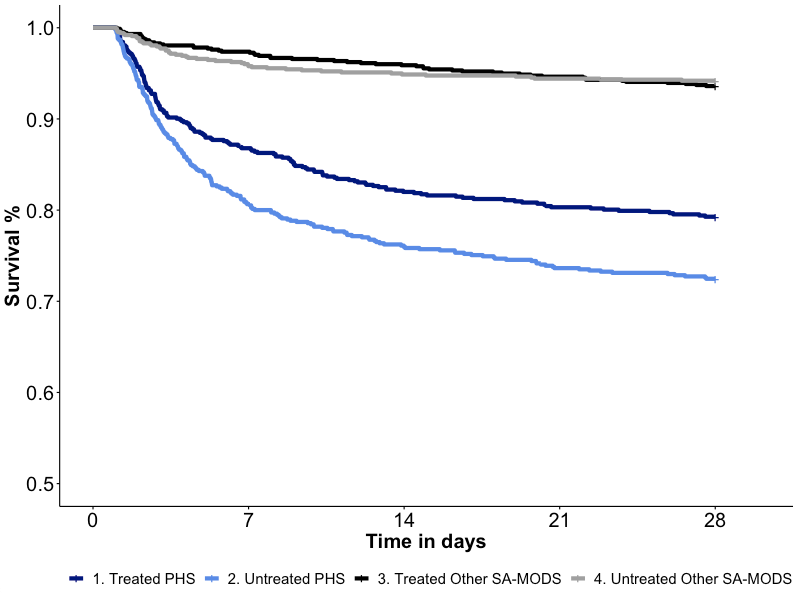

In the Cox regression analysis, there was a difference in the mortality of treated vs. untreated PHS patients (log rank test, p=0.002) but not in the SA-MODS patients (p=0.6)

Abbreviations: *MODS*, multiple organ dysfunction syndrome

**Additional file - Figure 6**. Covariate balance before and after propensity score matching in patients in the albumin analysis.

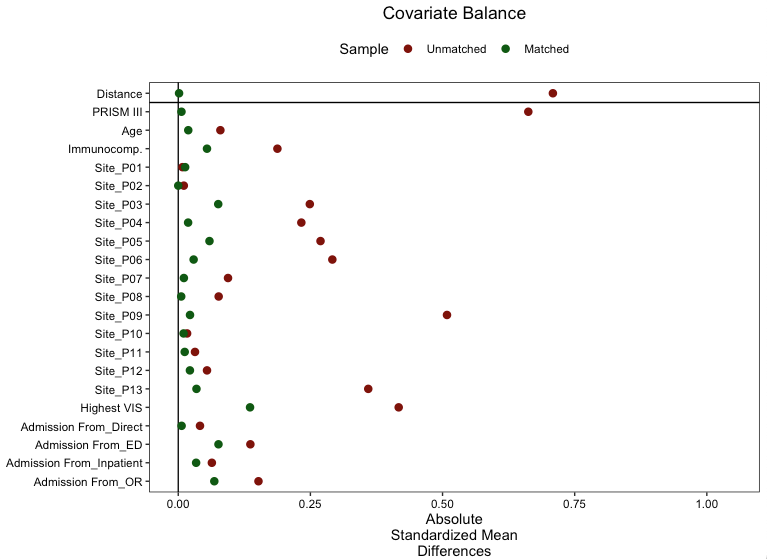

Abbreviations: *PRISM III*, Pediatric risk of mortality version 3; *Immunocomp.*, immunocompromised comorbidity.

**Additional file - Figure 7**. Kaplan-Meier survival curve* for patients with the *persistent hypoxemia and shock* phenotype and those with *other sepsis-associated-MODS* who were treated or not with albumin.

In the Cox regression analysis, there was a difference in the mortality of treated vs. untreated PHS patients (log rank test, p=0.006) but not in the SA-MODS patients (p=0.1)
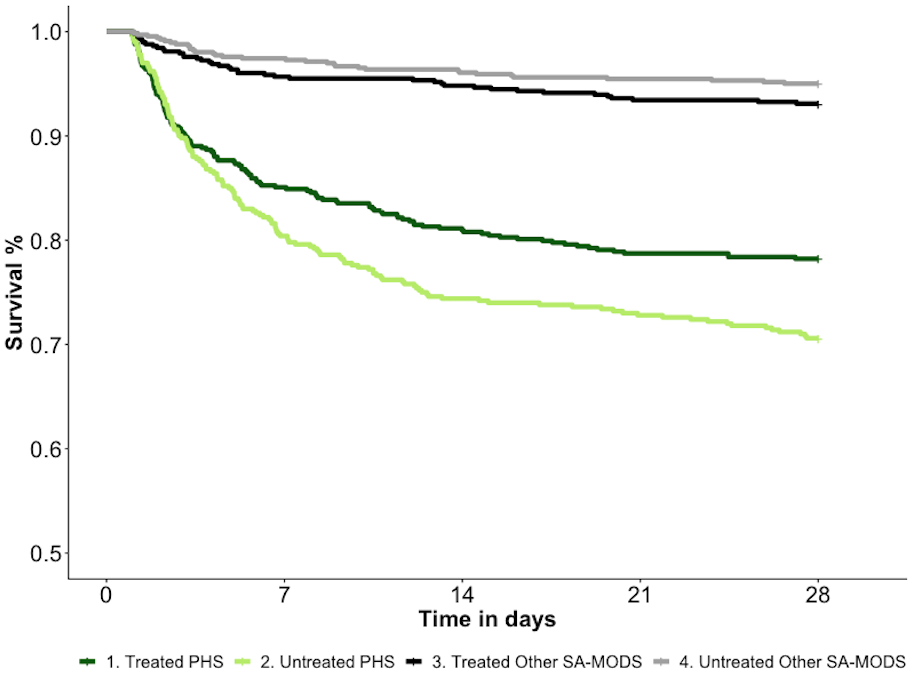

Abbreviations: *MODS*, multiple organ dysfunction syndrome; *PHS*, persistent hypoxemia and shock phenotype; *SA-MODS*, sepsis-associated MODS.

**Additional file - Table 8**. Distribution of patients with septic shock, moderate-to-severe pediatric ARDS, and in top tertile of the mean pSOFA score across the *persistent hypoxemia and shock* phenotype and patients with other sepsis-associated MODS in the first 72 hours of admission.

| Clinical Phenotype | Septic shock^a^ (n=5,793) | Moderate-to-severe pediatric ARDS^b^ (n=5,048) | Mean pSOFA in top tertile^c^ (n=5,059) |
| --- | --- | --- | --- |
| Persistent hypoxemia and shock, No. (%) | 3,209 (55.4) | 2,707 (53.6) | 3,150 (62.3) |
| Other sepsis-associated MODS, No. (%) | 2,585 (44.6) | 2,341 (46.4) | 1,909 (37.7) |

^a^Septic shock defined as requiring any vasoactive drip in the first 72 hours.

^b^Moderate-to-severe pediatric ARDS based on the hypoxemia criteria of the Pediatric Acute Lung Injury Consensus Conference (11): oxygenation index ≥ 8 or oxygen saturation index ≥ 7.5 in the first 72 hours.

^c^Mean pSOFA score cutoff for top tertile >6.25 in the first 72 hours.

Abbreviations: *ARDS,* acute respiratory distress syndrome; *MODS*, multiple organ dysfunction syndrome; *pSOFA*, pediatric sequential organ failure assessment.

**Additional file - Table 9**. Distribution of patients with the *persistent hypoxemia and shock* phenotype across age groups.

| Age group | Persistent Hypoxemia and Shock, No. (%) | p-value |
| --- | --- | --- |
| Less than 1 month (n= 611) | 272 (44.5) | <0.001 |
| 1 to 11 months (n= 2,677) | 969 (36.2) |  |
| 12 to 23 months (n= 1751) | 542 (31.0) |  |
| 2 to 4 years (n= 2,632) | 800 (30.4) |  |
| 5 to 11 years (n= 3,904) | 1,093 (28.0) |  |
| 12 years to 17 years (n= 3,671) | 1,160 (31.6) |  |
